# The Geography of Chronic Pain in the United States and Canada

**DOI:** 10.1101/2021.09.15.21263635

**Authors:** Anna Zajacova, Jinhyung Lee, Hanna Grol-Prokopczyk

## Abstract

Our understanding of population pain epidemiology is largely based on national-level analyses. This focus, however, neglects potential cross-national, and especially sub-national, geographic variations in pain, even though geographic comparisons could shed new light on factors that drive or protect against pain. This article presents the first comparative analysis of pain in the U.S. and Canada, comparing the countries in aggregate and analyzing variation across states and provinces. Analyses are based on cross-sectional data collected in 2020 from 2,124 U.S. and 2,110 Canadian adults 18 years and older. Our pain measure is a product of pain frequency and pain-related interference with daily activities. We use regression and decomposition methods to link socioeconomic characteristics and pain, and inverse-distance weighting spatial interpolation to map pain scores. We find significantly and substantially higher pain in the U.S. than in Canada. The difference is accounted for by Americans’ lower economic wellbeing. Additionally, we find variation in pain within countries; the variation is statistically significant across U.S. states. Further, we identify nine hotspot states in the Deep South, Appalachia, and the West where respondents have significantly higher pain than those in the rest of the U.S. or Canada. This excess pain is partly attributable to economic distress, but a large part remains unexplained; we speculate that it may reflect the sociopolitical context of the hotspot states. Overall, our findings identify areas with high need for pain prevention and management; they also other scholars to consider geographic factors as important contributors to population pain.

---

Our understanding of the epidemiology of chronic pain remains incomplete. One critical gap is the near absence of research on the geographic distribution of pain, especially at sub-national but also cross-national levels. Using unique cross-sectional data collected in 2020 from U.S. and Canadian respondents, we answer two key questions: (1) How is chronic pain geographically distributed in the U.S. and Canada, both with respect to the two countries in aggregate and within each country across states and provinces? And (2) how do population characteristics explain observed geographic differences in pain scores? Further, our analyses identify pain “hotspots,” examine predictors of excess pain in the hotspots, and, more broadly, invite future research to consider geographic factors as contributors to population pain.

The U.S. and Canada share many characteristics: Both are wealthy economies with highly educated, predominantly English-speaking, diverse populations and, according to most if not all classifications, share the “liberal” welfare-state regime designation [2; 39]. At the same time, they differ in important ways. Canada has lower poverty rates, less socioeconomic inequality, a stronger social safety net, and, unlike the U.S., a universal health care system [41;48]. Perhaps due to these differences, studies often—although not always—find longer lifespans and better health in Canada [14; 17]. However, no study to date has compared pain between these neighboring countries. Our U.S.-Canada comparison offers insights about social factors shaping pain at the national level.

Neither the U.S. nor Canada, however, is a monolithic entity. Both countries comprise subnational units–-states and provinces—that are increasingly heterogeneous with respect to health determinants [16; 31]. These include socioeconomic factors at the individual level, and specific policies as well as overall policy orientations at the macro level [13; 16]. Correspondingly, states and provinces vary substantially in health and mortality [27; 32; 52]; the variation is increasing over time [57]. For instance, age-standardized prevalence of disability, an important (often pain-related) dimension of health, is about 7% in Minnesota and North Dakota but over 13% in Kentucky or Mississippi [34]. Life expectancy also ranges widely across states and provinces, with a 6-year gap between the most and least healthy U.S. states [1], and a 3-year gap across Canadian provinces [49] -- or an 11-year gap, with the inclusion of the Northern territory of Nunavut [50]. Yet, research has nearly uniformly neglected subnational variability in pain and/or its predictors [13], as reflected by the absence of studies on pain prevalence across states and provinces.

Our study provides the first detailed examination of pain’s geographic distribution both across and within the U.S. and Canada, using a relatively fine-grained pain measure that combines pain frequency and pain-related interference with daily activities. Our findings identify hotspot areas with a particularly high need for pain prevention and treatment; clarify the role of demographic and socioeconomic population characteristics in explaining cross- and within-country differences; and pave the road for future studies of contextual factors that may help explain excess pain in hotspot regions.

## METHODS

### Data

We use the Recovery and Resilience COVID-19 data [51]. This cross-sectional dataset was developed by an interdisciplinary team of social science investigators at the University of Western Ontario and administered by Leger Opinion in the U.S. and Canada in August of 2020 [51]. The aim of the survey was to assess a wide range of sociopolitical, economic, and other conditions in the summer of 2020 in the two countries. The online survey was completed by 2,124 U.S. and 2,110 Canadian respondents aged 18 and older who were a part of an ongoing Leger Opinion Panel. The percentage of the panel respondents invited to participate in the survey who responded was 17% in Canada and 25% in the U.S. A small number of respondents were ineligible due to their age (below 18) or their responses were eliminated due to low quality; the final response rate was 13% in Canada and 19% in the U.S. (We briefly discuss the response rate in the discussion section.) The samples were designed to be nationally representative of age, gender, and Census region in the U.S. and provinces in Canada, and sampling weights were provided by Leger to correct for over- and under-sampling; the weighted sample is representative of the population with respect to these three characteristics. The survey was approved by the Ethics Board of the University of Western Ontario (Project ID 116046).

### Variables

Pain was assessed with two questions. First, respondents were asked: “in the past 30 days… How often have you experienced pain?” The response options were never or almost never, rarely, sometimes, often, almost always, and always (coded as 0-5). Respondents who chose an option other than ‘never or almost never’ were then asked: “how much did the pain interfere with your general activity like work or household chores?” This item was assessed on an 11-point scale from ‘did not interfere’ to ‘completely interfered.’ The pain score was created as the arithmetic product of these two variables (frequency and interference). This merged score yielded a scale from 0 to 55. The score of 0 is for respondents who reported they experienced pain “never or almost never,” while 55 is for those reporting that they ‘always’ had pain that ‘completely interfered’ with their everyday activities. This variable had a right-skewed distribution, which we found to be best modeled using negative binomial regressions. We also dichotomized the full scale for sensitivity analyses using 2 thresholds: a pain score of at least 10 and a pain score of at least 20. The threshold of 10 corresponds to a less stringent pain definition that included even relatively infrequent and low-interference pain, such that 42.5% of the combined U.S./Canada target population reported pain above that threshold. The threshold of 20 is a more stringent pain definition reflecting either more frequent or higher-impact pain, reported by 21.4% of the target population.

State and province of residence was reported by the respondents; U.S. respondents chose from a list of states and Canadian respondents from a list of provinces.

Age was collected using the following categories: 18-24, 25-34, 35-44, 45-54, 55-64, and 65+. We collapsed the middle categories in some analyses for parsimony, generating categories of 18-24, 25-44, 45-64, and 65+. These two specifications yielded substantively identical results. Gender was collected as male (reference), female, or other. Race was recoded to white (reference), Black, Hispanic, Asian, and other. Immigrant status was a dichotomy of native-born (reference) versus immigrant. Marital status was coded as married (reference, includes common-law), previously married, and never married. Respondents also reported whether they had children or not (reference). Education was coded as high school or less, some postsecondary education, associate degree or equivalent, and bachelor’s degree or more (reference). Main activity status included the categories employed (reference), retired, unemployed, disabled, and other. Finally, respondents were asked about their family income and financial hardships due to COVID-19, which can be viewed as indicators of longer-term and short-term economic well-being or stress. Family income was categorized as $0-29,000 (reference); $30-59,000, $60-89,000, 90-149,000, and $150,000 or more. The same categories were used in the U.S. and in Canada, without identifying dollars as U.S. or Canadian. We did not convert the currency because we felt that these five broad categories captured the relative income standing in each country sufficiently well. Financial hardship was measured with the question “Are you feeling financial hardship due to COVID-19?” and coded as no hardship (reference), little or slight hardship, some hardship, or serious hardship.

### Approach

Our approach had three major analytic steps. We first calculated univariate and bivariate (U.S. versus Canada) descriptive statistics. Specifically, we estimated weighted pain scores in the aggregate and for each sociodemographic group in each country; we also tested whether the mean pain scores differed between the two countries, using group-specific bivariate negative binomial regression models on a country indicator (Table 1). We also estimated the distribution of all covariates in each country, and tested for differences in the distributions between the two countries using design-adjusted F-tests (Supplemental Table S1).

**Table 1.** Mean pain scores in the U.S. and Canada for the total sample and in each subgroup.

|  |  | U.S. |  | Canada | p-value from Wald test of difference |
| --- | --- | --- | --- | --- | --- |
| Pain score in full sample | 12.5 | (12.0, 13.1) | 10.7 | (10.2, 11.3) | <.001 |
| Pain score, by age |  |  |  |  |  |
| 18-24 | 8.0 | (6.6, 9.3) | 8.0 | (6.9, 9.1) | .969 |
| 25-44 | 13.6 | (12.4, 14.8) | 10.1 | (9.3, 10.9) | <.001 |
| 45-64 | 13.8 | (12.6, 14.9) | 11.5 | (10.5, 12.4) | .002 |
| 65+ | 11.2 | (9.8, 12.6) | 11.8 | (10.5, 13.0) | .553 |
| Pain score, by gender |  |  |  |  |  |
| Men | 11.7 | (10.8, 12.6) | 9.7 | (8.9, 10.4) | .001 |
| Women | 13.4 | (12.5, 14.3) | 11.7 | (10.9, 12.4) | .004 |
| Pain score, by race/ethnicity |  |  |  |  |  |
| Non-Hispanic white | 13.1 | (12.3, 13.9) | 11.2 | (10.5, 11.8) | <.001 |
| Non-white | 10.7 | (9.6, 11.9) | 9.4 | (8.4, 10.3) | .071 |
| Pain score, by immigrant status |  |  |  |  |  |
| Not immigrant | 12.9 | (12.2, 13.5) | 11.0 | (10.4, 11.6) | <.001 |
| Immigrant | 8.5 | (6.5, 10.6) | 9.4 | (8.4, 10.4) | .474 |
| Pain score, by education |  |  |  |  |  |
| High school or less | 13.1 | (11.6, 14.6) | 12.9 | (11.5, 14.2) | .819 |
| Some postsecondary | 14.6 | (13.1, 16.0) | 11.7 | (10.4, 13.0) | .004 |
| AA or VTC | 13.5 | (11.6, 15.4) | 13.5 | (12.2, 14.9) | .984 |
| BA or higher | 10.9 | (10.0, 11.8) | 8.1 | (7.5, 8.7) | <.001 |
| Pain score, by main activity |  |  |  |  |  |
| Employed | 11.2 | (10.4, 12.1) | 9.4 | (8.7, 10.0) | <.001 |
| Retired | 12.1 | (10.5, 13.7) | 11.9 | (10.7, 13.1) | .874 |
| Unemployed | 13.0 | (10.7, 15.3) | 11.5 | (9.3, 13.6) | .335 |
| Disabled | 25.5 | (22.6, 28.4) | 32.3 | (28.0, 36.5) | .008 |
| Other | 11.5 | (9.9, 13.0) | 9.1 | (7.9, 10.3) | .020 |
| Pain score, by family income |  |  |  |  |  |
| 0-29k | 15.7 | (14.4, 16.9) | 14.7 | (13.2, 16.2) | .352 |
| 30-59k | 11.7 | (10.5, 12.9) | 12.0 | (10.8, 13.1) | .751 |
| 60-89k | 10.8 | (9.3, 12.4) | 9.8 | (8.8, 10.8) | .249 |
| 90k or more | 10.2 | (9.1, 11.4) | 8.2 | (7.5, 8.9) | .002 |
| Pain score, by financial hardship<sup>1</sup> |  |  |  |  |  |
| No hardship | 9.6 | (8.7, 10.6) | 8.0 | (7.4, 8.7) | .005 |
| Slight/little hardship | 12.3 | (11.3, 13.2) | 11.0 | (10.2, 11.8) | .041 |
| Some hardship | 14.7 | (12.8, 16.6) | 16.4 | (14.4, 18.3) | .232 |
| Serious hardship | 22.2 | (19.3, 25.2) | 19.4 | (15.9, 22.9) | .229 |
| Dichotomized pain score |  |  |  |  |  |
| Pain ≥ 10 | 45.4% | (42.9%, 47.9%) | 39.6% | (37.5%, 41.8%) | <.001 |
| Pain ≥ 20 | 23.9% | (21.8%, 25.9%) | 19.0% | (17.3%, 20.7%) | <.001 |
| Pain ≥ 30 | 13.0% | (11.4%, 14.6%) | 8.8% | (7.6%, 10.1%) | <.001 |

Weighted means and 95% confidence intervals. Pain is a continuous variable with 0-55 range. N=4,162. The p-values are from Wald tests for a U.S. dummy (versus Canada) in weighted negative binomial bivariate regression of pain estimated separately for each group.
<sup>1</sup> Financial hardship due to COVID-19

In our second major analytic step, we analyzed differences between U.S. and Canada in aggregate, using two complementary approaches: nonlinear decomposition (Table 2) and a series of regression models (Supplemental Table S2). The Oaxaca-Blinder nonlinear decomposition is a widely-used econometric method that quantifies how much of the difference in pain prevalence between the U.S. and Canada is due to different population characteristics (composition) or different coefficient effects (also referred to unexplained part) [3; 40]. More formally, the observed difference in pain prevalence 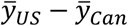, where 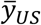 is the mean pain level in the U.S. and 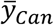 is the mean pain level in Canada, is defined as 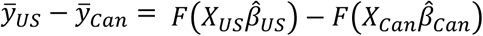, where the *X*_*US*_ and *X*_*Can*_ are vectors of observed covariates in the U.S. and Canada, respectively. Their associated vectors of coefficients 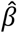 s are estimated using a negative binomial model appropriate to the skewed positive distribution of the pain scores, and *F*() is the cumulative distribution function of the negative binomial distribution. The term 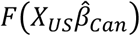 can be added and then subtracted to obtain: 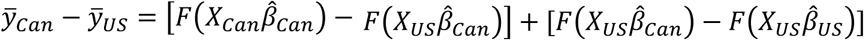. The first bracket captures the gap between the two countries due to differences in population characteristics while the second bracket captures the part due to differences in coefficients. We used the *mvdcmp* extension in Stata for decomposition [43], combined with the new utility for grouping individual covariates for detailed decomposition, *mvdcmpgroup* (Powers 2020, personal communication). The effects of categorical variables in this approach are normalized as deviations from the grand mean, which enables calculation of effects for all levels, and yields results that are identical regardless of which category is the reference [22]. In the supplement, we also show the more widely used, though more constrained, approach to examining the country-level differences: a series of weighted negative binomial regression models with an indicator for the U.S., net of different sets of covariates. The drawback in the regression models is that the effect of all covariates is constrained to be equal in both countries, effectively equivalent to forcing the coefficient effect (unexplained part) in the decomposition to be equal to zero.

**Table 2.** Nonlinear decomposition of the pain score difference between US and Canada due to differences in population composition versus differences in coefficient effects.

|  | Due to composition differences (%) | Due to coefficient differences<sup>1</sup> (%) |
| --- | --- | --- |
| <u>Total decomposition</u> | 82.2*** | 17.8 |
| <u>Detailed</u> |  |  |
| Age and sex | 5.0** | 4.4 |
| Race and immigrant status | 26.3** | 16.2 |
| Marital and parent status | 3.6 | -1.9 |
| Education | -0.8 | 9.9 |
| Economic factors | 48.1*** | 5.0 |

\*p<.05, \*\*p<.01, \*\*\*p<.001
The approach decomposes the difference in mean pain score in the U.S. (12.5) versus in Canada (10.7) into a part due to differences in population characteristics (composition) and a part due to differences in coefficients (that is, the effects of the coefficients on pain). The group "Economic factors" includes main activity, family income, and financial hardship due to COVID-19.
<sup>1</sup> This part of the difference is also referred to as the unexplained part.

Our third major analytic step was to analyze the pain scores at the level of subnational units, that is, states in the U.S. and provinces in Canada. First, we mapped the geographic distribution of weighted mean pain scores in the U.S. and Canada (Figure 1). The inverse distance weighting (IDW) spatial interpolation technique [26] was used to estimate pain scores for unsampled locations using values from surrounding locations, thereby generating a continuous surface of weighted mean pain scores across the U.S. and Canada. Table 3 provides a different perspective: it lists weighted mean pain scores in each state/province with at least 15 respondents, ranked from highest to lowest score. Table 3 also lists estimated proportions of residents with pain scores ≥10 and ≥20, respectively, as robustness checks. These descriptive steps identified a set of hotspot states with the highest pain levels. We then tested whether the pain scores vary significantly across all subnational units via a likelihood ratio F-tests of joint significance (Supplemental Table S3). Finally, we used nonlinear decomposition (Table 4) and negative binomial regression models (Supplemental Table S4), akin to the methods in Tables 2 and S2, to examine the sources of the excess pain in the hotspot areas.

**Table 3.**
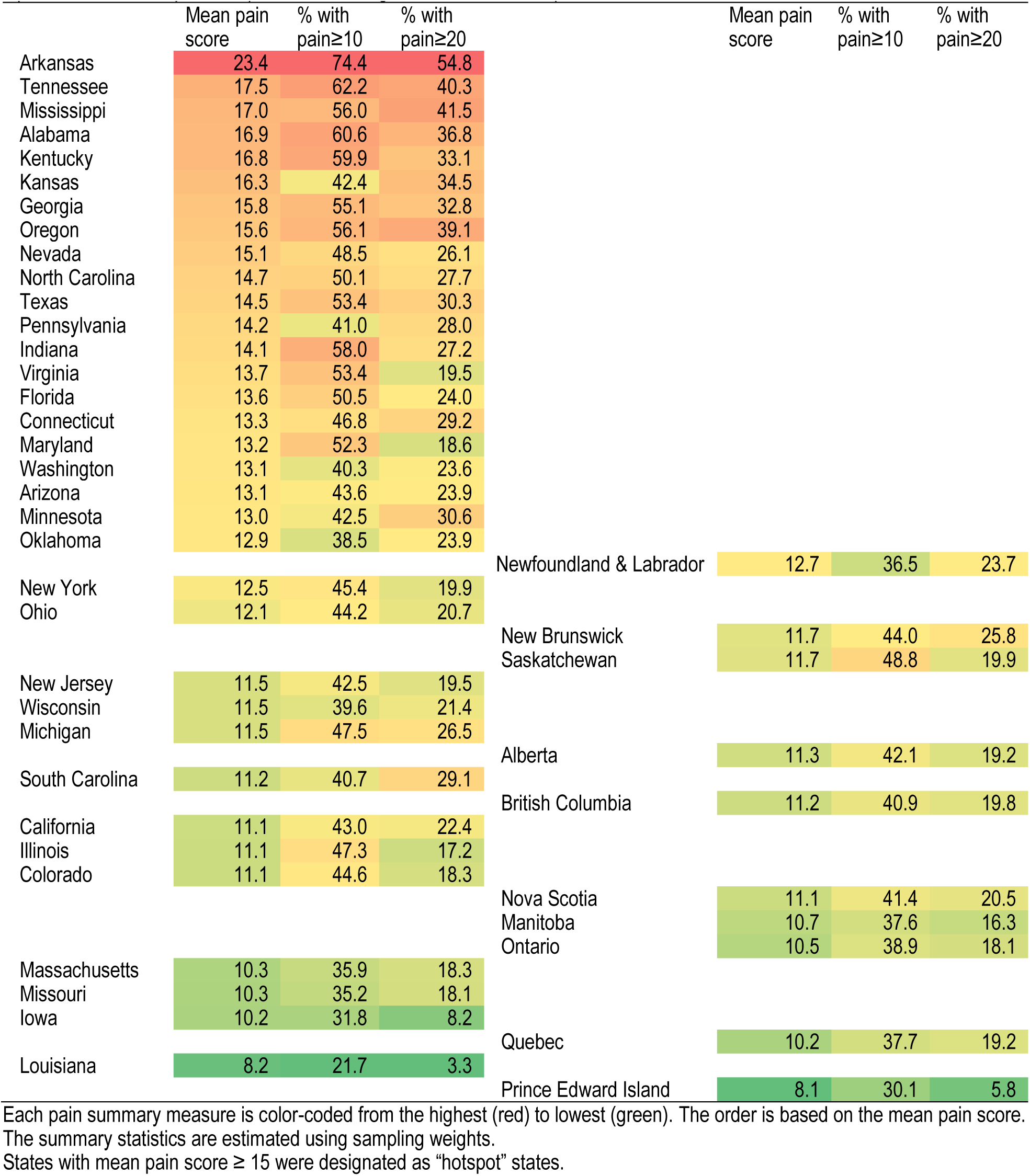
Weighted mean pain score and proportions with pain scores ≥ 10 and ≥20, by U.S. state and Canadian province (with 15 or more respondents), ranked from highest to lowest mean pain score

**Table 4.** Nonlinear decomposition of the pain score difference between hotspot states and all other areas due to differences in population composition versus differences in coefficient effects.

|  | Due to composition differences (%) | Due to coefficient differences<sup>1</sup> (%) |
| --- | --- | --- |
| <u>Total decomposition</u> | 41.8** | 58.2** |
| <u>Detailed</u> |  |  |
| Age and sex | 2.5 | 8.5 |
| Race and immigrant status | 8.6 | 57.1 |
| Marital and parent status | -0.4 | 6.6 |
| Education | -2.6 | 2.5 |
| Economic factors | 33.7** | -0.5 |

\*p<.05, \*\*p<.01, \*\*\*p<.001
States with weighted mean pain score $\geq 15$ were designated as “hotspot” states.
The approach decomposes the difference in mean pain score in the 9 hotspot states versus all other subnational units into a part due to differences in population characteristics (composition) and due to differences in coefficients (that is, the effects of the coefficients on pain). The group “Economic factors” includes main activity, family income, and financial hardship variables.
<sup>1</sup> This part of the difference is also referred to as the unexplained part.

**Figure 1.**
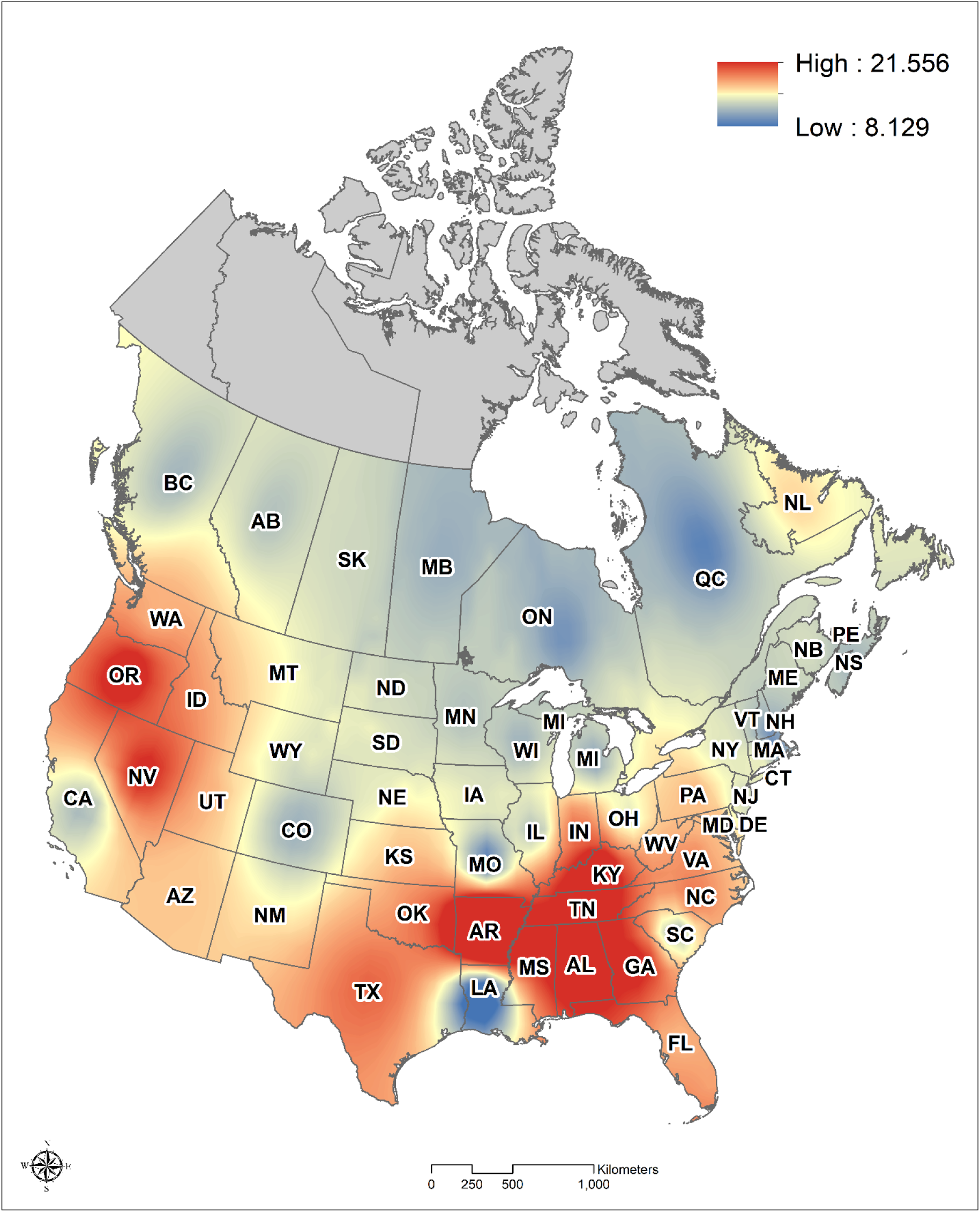
Mean pain scores in U.S. states and Canadian provinces. Map visualizes weighted mean pain scores in each state and province.

All analyses were weighted. Data cleaning and most analyses were conducted in Stata 17; the mapping was done in ArcMap 10.8.1.

## RESULTS

Table 1 summarizes weighted mean pain scores in the U.S. and Canada, both within the full sample and in population subgroups. In the total sample, the mean pain score was 12.5 (95%CI 12.0, 13.1) in the U.S. and 10.7 (95%CI 10.2, 11.3) in Canada, a statistically significant difference (p<.001). We also dichotomized the pain score at three cutpoints. For each of these specifications, again, pain was significantly higher in the U.S.: Americans had 5.8 percentage points (pp) greater change of pain≥10, 4.9 pp greater chance of pain≥20, and 4.2 pp greater chance of pain≥30.

In every subgroup, pain was either statistically significantly higher in the U.S., or the between-country difference was not significant. The groups with the highest average pain scores in both countries were those who reported serious financial hardship due to COVID-19 (with average pain scores of 22.2 in the U.S. [95%CI 19.3, 25.2] and 19.4 in Canada [95% CI 15.9, 22.9]), and respondents who described their main activity status as “disabled” (25.5 in the U.S. [95%CI 22.6, 28.4] and 32.3 in Canada [95% CI 28.0, 36.5]). Further, we note an interesting general pattern whereby the U.S. pain ‘excess’ vis-à-vis Canada tends to be most pronounced among more socially advantaged groups. That is, the pain ‘excess’ in the U.S. versus in Canada is statistically significant for white, non-immigrant, college-educated, employed, high-income, and no/little financial hardship groups. The differences are smaller and not significant among those who are non-white, immigrant, low-educated or recipients of a subbaccalaureate degree like associate degree or vocational/technical certificate, lower income, and experiencing some/serious financial hardship due to COVID-19.

Supplemental Table S1 summarizes the distribution all covariates in the U.S. and Canadian samples and shows p-values from tests for differences in the distribution of each covariate between the countries. The two samples differ with respect to several demographic characteristics, in particular race/ethnicity and immigrant status, but also gender. The U.S. adult population comprises more Black and Hispanic respondents than the Canadian population, where Asian Canadian and ‘other’ groups predominate among non-white groups. Additionally, the U.S. sample includes 7.5% immigrant adults, compared to 19.3% in Canada. Race/ethnicity and immigrant status matter greatly in analyses of pain because immigrants report less pain than native-born adults, at least in the U.S. [18; 58;59]. There are important racial/ethnic differences in pain prevalence as well [36; 42; 60], with adults of Asian heritage tending to report particularly low pain prevalence [25; 35]. With respect to gender, pain differences are well known; women report greater pain prevalence, severity, and interference, compared with men [4; 44; 54]. Thus, inter-country differences in race/ethnicity, immigrant status, and gender may shape overall country differences in pain. The U.S. and Canadian samples also differ in socioeconomic status indicators. The U.S. has significantly lower educational attainment and family income, and U.S. respondents reported significantly greater financial hardship due to COVID-19 (all p<.001). Given that socioeconomic status is one of the most important social factors in pain [21; 29; 45], we next examine how these differences impact pain in both countries.

Our second step was to analyze the sources of the pain score difference between the U.S. and Canada. Table 2 shows the proportion of the observed pain score difference which can be attributed to differences in composition (population characteristics) versus differences in coefficient effects (unexplained part). The total decomposition shows that 82.2% of the gap in the mean pain scores between the two countries is due to differences in composition (characteristics) while 17.8% is unexplained, that is, due to differences in coefficient effects. The detailed decomposition shows how the covariates, grouped for parsimony, add up to these total percentages. Nearly half of the compositional difference is from economic factors, i.e., main activity, family income, and financial hardship (48.1%, p<.001). As we summarized in Table S1, U.S. respondents have lower family income and greater financial hardship than their Canadian counterparts. The second major source is the different race and immigrant status composition (26.3%, p<.01). This is also unsurprising given the high proportion of Canadians who are immigrants or Asian Canadians, characteristics associated with particularly low chronic pain prevalence as noted in the prior paragraph. None of the coefficient differences, whether total or specific to any group of covariates, is a statistically significant contributor to explaining the U.S. excess in pain.

Supplemental Table S2 corroborates this picture with a set of regression models of pain scores as a function of the U.S. indicator and covariates. This set of models effectively constrains the effect of all covariates to be equal in the U.S. and Canada since interaction terms are not included; however, this constraint is reasonable since the decomposition analysis found no statistically significant differences in the coefficient effects. The results show that the incidence rate ratio (IRR) of pain is 1.17 in the U.S. relative to Canada, whether or not we control for age and gender distribution (Models 1 and 2, both p<.001). Race and immigrant status attenuate the U.S. pain disadvantage somewhat (IRR=1.14, p<.001 in Model 3) while marital status and children are not significant predictors and correspondingly change the U.S.-Canada difference little (IRR=1.13, p<.01 in Model 4). Just as the decomposition analysis showed, economic indicators, included in Models 5-8, are significant predictors of pain and jointly also explain the U.S. excess pain, which is no longer significant in Models 6 or 8 (IRR=1.07 and 1.05, respectively; p>.05 in both cases). Education is a suppressor: controlling for this covariate actually makes the U.S. disadvantage significant, with or without inclusion of economic indicators (IRR=1.08, p<.05 in Model 10). In sum, the decomposition and regression analyses show that most although not all of the higher pain score in the U.S. compared with Canada is due to (a) the greater economic stress among U.S. adults, and (b) racial/immigrant compositional differences between the two countries.

The analyses thus far treated both countries as monolithic units, when in reality they comprise potentially heterogeneous subnational units. Therefore, we next examine how pain varies within each country across states and provinces. Figure 1 shows a map of weighted mean pain scores in the U.S. and Canada. The map highlights the relatively low average pain scores across most of Canada (blue hue) except Newfoundland and Labrador, as well as across much of the U.S. Midwest and Northeast. The figure also shows that pain is substantially higher in much of the U.S. Deep South and parts of the Appalachia, as well as select areas of the West and Northwest (particularly Oregon and Nevada), as the red hue indicates.

We also quantified the mean pain levels by state or province. Table 3 shows the mean pain scores, as well as the proportion of respondents whose pain scores exceed 10 points and 20 points, respectively, in each state or province with at least 15 respondents, ordered from highest to lowest mean scores. The left panel in the table lists U.S. states; the right panel lists Canadian provinces. The average pain scores in U.S. states range from about 10 points in Iowa, Missouri, Massachusetts, and --surprisingly, given its overall poor health [53]--Louisiana, to 17 or above in Arkansas, Tennessee, and Mississippi. The proportions with pain (using either the 10- or 20-point thresholds to dichotomize the pain scores) yield generally comparable ranking of states. The range of pain scores across Canadian provinces, in the right-hand side panel, is substantially smaller. The pain scores range from about 8-10 in Prince Edward Island, Quebec, and Ontario, to less than 13 in Newfoundland and Labrador, New Brunswick, and Saskatchewan. All the Canadian provinces have pain scores on par with the lower half of U.S. states. In other words, about half of U.S. states have pain scores higher than *any* province in Canada.

Is the subnational heterogeneity statistically significant? Supplemental Table S3 answers this question by testing the joint effects of U.S. states, Canadian provinces, and all subnational units jointly, in negative binomial regression models of pain net of different covariate sets. The variation is significant in the null model (p<.05) and marginally when we control for age and sex (p<.10) when we analyzed all subnational units, that is, states and provinces together. When we control for additional characteristics, however, the joint effect of subnational units is not significant. The variation is not statistically significant for any U.S. and Canada-specific models, perhaps because of the relatively modest sample sizes for most subnational units (Supplemental Table S4 lists the sample sizes for each state and province).

However, although the variability of all subnational units is only marginally significant net of age and sex, the high level of pain in the states with the highest pain scores warrants further examination. We designated states with average pain score ≥ 15 as “hotspot” states. The hotspot states included Arkansas, Tennessee, Mississippi, Alabama, Kentucky, Kansas, Georgia, Oregon, and Nevada, states clustered in the Deep South, parts of Appalachia and the Northwest. We next tested whether the hotspot states differ significantly from the rest of the subnational units, using nonlinear decomposition and also regression models.

The results from negative binomial regression models of pain net of an indicator for the hotspot states (Arkansas, Tennessee, Mississippi, Alabama, Kentucky, Kansas, Georgia, Oregon, and Nevada) versus all other subnational units, net of all covariate sets, are shown in Supplemental Table S5. The Table shows that the IRR of pain in hotspot states is 1.45 (p<.001) higher relative to all other units in unadjusted Model 1. About a third of this excess pain appears to be due to economic factors, especially family income. Controlling for these covariates (Models 6-9) attenuates the U.S. excess by about one third (IRR=1.29, p<.001 in Model 9). However, even after controlling for all covariates, the hotspot states still have significantly higher pain than other subnational units (IRR=1.30, p<.001), highlighting that much of the difference in pain levels remains unexplained. We also note that the difference between the hotspot states versus all other units is considerably greater than the differences between the U.S. and Canada, which highlights that the within-country variation can be, and in our case is, greater than the cross-country difference.

A complementary portrait of the sources of the hotspots’ excess pain is in Table 4, which shows results from a nonlinear decomposition in which we decompose the difference in pain in the hotspot states versus all other states and provinces. The results show that 41.8% of the pain excess in the hotspot states is due to differences in the distribution of the observed population characteristics between these hotspots and other states/provinces. The majority of this explained (compositional) part is economic factors (33.7%, p<.01), which is similar to the result from the regression models in Supplemental Table S5. Over 58% of the hotspot excess, however, is ‘unexplained,’ that is, due to different effects of observed characteristics on pain or a residual difference in the intercepts due to factors not included in the study. Thus, both regression and decomposition perspectives suggest that the pain excess in the hotspot areas is due partly to economic disadvantage of the hotspot-area residents, but in large part to factors beyond those we included in the analysis.

## DISCUSSION

The Federal Pain Research Strategy has declared that the “greatest near-term value” research priority to better understand pain disparities is to “better define the epidemiology of pain in disparate populations,” and the “most impactful top priority” is to investigate mechanisms, including social mechanisms, that contribute to group differences in chronic pain [20, p. 18]. Our analysis contributes directly to these priorities. Using a new international survey that allowed us to combine pain frequency and interferences with daily activities, we compared population pain levels in the United States and Canada at the aggregate national level and tested social correlates of observed differences. Further, we examined geographic variation in pain *across* states and provinces, identified hotspots with particularly high pain levels, and checked whether these hotspots are a function of sociodemographic characteristics within those areas.

We found that pain is significantly higher on average in the U.S. than in Canada. This was the case regardless of whether we used a continuous pain score for the comparison, or a dichotomized scale. The latter showed a 4-6 percentage point higher pain burden in the U.S. across different scale cutpoints. This difference is clinically meaningful and translates to roughly 10 million extra U.S. adults experiencing pain compared to what it would be at the Canadian levels. While ours is the first comparative study of pain in these two countries, our findings corroborate relevant comparative analyses, which found worse health and higher mortality in the U.S. than in Canada [5; 14; 17].

The higher pain burden in the U.S. versus Canada appears to be a function of the worse economic conditions of U.S. adults, including greater likelihood of low income and financial hardship. The importance of these factors is unsurprising, as the strong impact of economic distress on physical pain has been well documented [6; 23;29; 55; 56]. Our counterfactual decomposition suggests that if family income and financial hardship were equal in the U.S. and Canada, there might be no difference in the pain burden between the two countries. Additionally, the decomposition showed that the effects of socioeconomic factors on pain were comparable in the U.S. and Canada. This is an important finding because it confirms the link between socioeconomic factors and pain burden at a national level, which in turn contributes to the foundational evidence base regarding the importance of social roots of pain.

However, the pain patterns were more complex at the subnational level. We found variation across states/provinces, although it was statistically significant only for all subnational units together, not when U.S. states and Canadian provinces were examined separately. In Canada, the pain scores ranged from about 8-10 in Quebec, Ontario, and Prince Edward Island, to around 12-13 in the Atlantic provinces of Newfoundland and Labrador and New Brunswick; the Prairies and the Canadian West were roughly in the middle. This general geographic pattern fits the findings of the only prior study estimating province-specific pain prevalence, which used a large nationally-representative Canadian health survey [47].

The pain scores ranged more widely in the U.S., from about 10 in Iowa, Missouri, and Massachusetts, to over 23 in Arkansas. Overall, about half of U.S. states had pain burden within the range observed across Canadian provinces, while the other half had more pain than found in *any* Canadian province. Indeed, some states had such high pain burden relative to all other sub-national units (pain score≥15) that we designated these states as pain “hotspots.” These comprise the primarily Southern and Appalachian states of Arkansas, Tennessee, Mississippi, Alabama, Kentucky, Georgia, as well as Kansas, and Western states of Oregon and Nevada. Jointly, these hotspot areas have a significantly and substantially greater pain burden than other states and provinces, about 45% higher incidence rate. There are no prior analyses of pain burden across U.S. states to compare our findings to; indeed, the majority of studies on U.S. population pain burden include no geographic indicators [7; 11; 15; 61]. However, several studies described U.S. pain by Census region and reported higher chronic pain in the South [37] or West [24; 59] and less in the Northeast [37; 58]. Our findings are generally consistent with these patterns, as well as with geographic patterns in other dimensions of health, which tends to note particularly high rates of mortality [32; 57], disability [34], and risk factors such as smoking [8] and obesity [9] in the Deep South and Appalachia states.

The absence of geographic variation in population pain is particularly surprising when contrasted with a much better understanding of variation in pain *treatment*, especially opioid use and misuse, across U.S. states and Canadian provinces. The hotspot areas we identified with respect to pain overlap with areas of high opioid use: Appalachia and the South [10], and states in the Pacific Northwest [28; 46]. Whether this overlap is coincidental, causal, or confounded by common causes, it requires further investigation, and we urge the collection and public-use dissemination of data that would enable scholars to examine the geography of chronic pain.

About 33-42% of the excess pain in the hotspot areas is due to differences in sociodemographic factors and economic conditions (with the lower bound of this range based on regression analyses, and the upper bound based on decomposition). The majority of the pain excess, however, is due to factors unobserved in our study. Such factors may include sociopolitical features of the hotspot states, such as health care system generosity, minimum wage floors, taxation, housing policies, environmental protection, and even structural sexism and racism, all of which are connected to population health [19; 30; 32]. These sociopolitical features reflect states’ overall policy orientations: For instance, Minnesota supports their residents’ health and wellbeing more than a state like Mississippi, and such policy differences are then powerfully reflected in the overall health and mortality of these states’ residents [33]. It is reasonable to expect that such state differences would also be reflected in pain burden. Other unobserved drivers of the excess pain in the hotspot areas could also be additional factors measured at the individual level, such as obesity or social support. However, it is important to remember that federal, state, or local contexts and policies are crucial in shaping individual circumstances, as well as their effect on health. Future research could consider the role of national or subnational policies (e.g., redistributive taxes such as the Earned Income Tax Credit, food stamp eligibility, Medicaid or other health insurance eligibility policies, and other social safety net features) in shaping pain, via mechanisms like income and financial hardship.

Our conclusions are limited by sample size and representativeness. While our total sample exceeds 4,000 (with over 2,000 respondents in each country), the subnational analyses, especially with respect to the 50 U.S. states, would ideally be based on a larger sample. Thus, our conclusions about any single state, especially outliers like Arkansas and Louisiana (both of which had only 21 respondents), must be viewed as provisional. Louisiana’s low pain burden was particularly unexpected due to its low position in health and longevity rankings [34; 38; 53]. We note that pain levels in Louisiana were not actually the lowest in our dataset, rather, states including South and North Dakota, Hawaii, and Utah had lower pain scores, as could be expected on the basis of their overall healthy profiles [34; 38]. However, they included fewer than 15 respondents, which we set *a priori* as a threshold for presenting state-specific findings.

The Recovery and Resilience COVID-19 survey’s low response rate, moreover, raises the possibility of biased selection into the sample. The distribution of sample characteristics indeed suggests that both the U.S. and Canadian samples undersampled disadvantaged respondents, such as those of low socioeconomic status (SES). The sampling weights adjust for age, sex, and region/province distribution, but not for SES or other characteristics. This suggests that our aggregate estimates of pain scores likely underestimate the true burden since the omitted disadvantaged adults would likely report higher pain [12; 59]. However, there is no reason to believe that the selection processes into the U.S. versus Canadian samples, or into samples in individual states and provinces, would differ systematically and bias the reported comparisons.

## Conclusion

Overall, population pain is higher in the U.S. than in Canada, largely due to the worse economic conditions among U.S. adults. While our associational cross-sectional study does not allow us to make policy recommendations, these findings suggest that easing Americans’ economic stress may, in addition to other benefits, lessen the pain burden experienced by the population. Additionally, pain variation across states and provinces was even larger than between the two countries. In particular, a portion of U.S. states in the Deep South, parts of Appalachia and the West had pain levels high enough to be designated as “hotspots.” We posit that state policy orientation and context may help explain the hotspots’ excess pain. Future analyses should draw on cross-national and sub-national variation in pain as fresh lens to uncovering the macro-to individual-level social roots of population pain.

## Data Availability

The data will be posted by November 2021 at a server location accessible to all interested researchers. The URL will be determined at that time and the information will be updated in the article pdf. Scholars may also after November 2021 for the URL.

## Acknowledgements

This study was supported in part by the National Institute on Aging (R01AG055481, R01AG06535101A1) and the Canadian Social Science and Humanities Research Council Insight Grant. The authors thank Cecilia Diaz Campo, Dr. Jason Winders, Dr. Autumn Knowlton, and Lindsay Finlay for their invaluable assistance with manuscript and analysis development, and Dr. Laura Stephenson for leading the development of the survey instrument and data collection.

All authors declare no conflict of interests.

## Supplement

**Supplemental Table S1.**
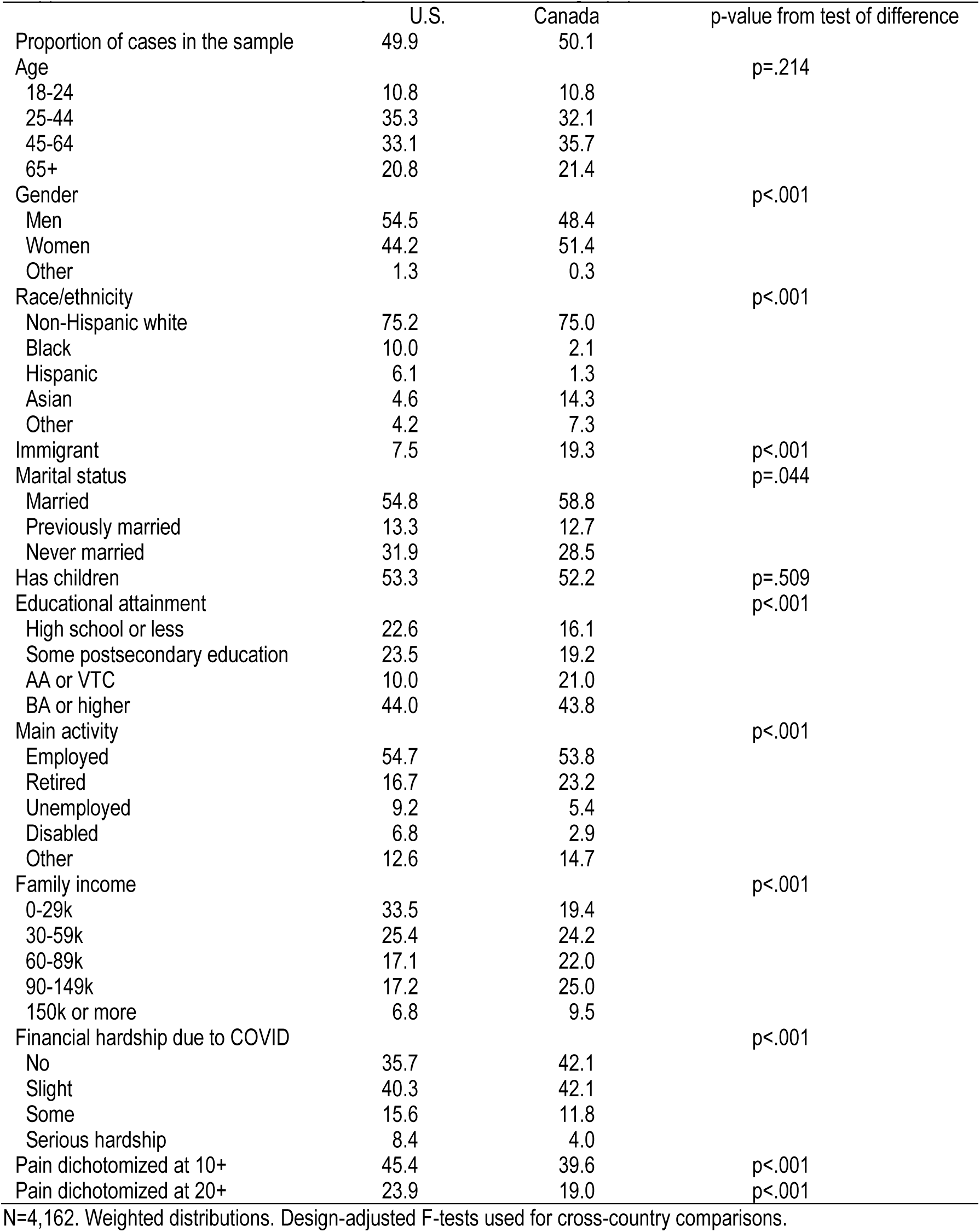
Distribution of key characteristics in the target population in the U.S. and Canada

**Supplemental Table S2.** Negative binomial regressions of pain score on country, net of covariates.

N=4,162. Weighted distributions. Design-adjusted F-tests used for cross-country comparisons.
|  |  |  |  |  |  |  |  |  |  |  |
| --- | --- | --- | --- | --- | --- | --- | --- | --- | --- | --- |
|  | (1) | (2) | (3) | (4) | (5) | (6) | (7) | (8) | (9) | (10) |
| U.S. (vs. Canada) | 1.17*** | 1.17*** | 1.14*** | 1.13** | 1.11** | 1.07 | 1.08* | 1.05 | 1.16*** | 1.08* |
| Age (18-24) |  |  |  |  |  |  |  |  |  |  |
| 25-44 |  | 1.50*** | 1.48*** | 1.43*** | 1.38*** | 1.40*** | 1.45*** | 1.37*** | 1.58*** | 1.43*** |
| 45-64 |  | 1.61*** | 1.53*** | 1.46*** | 1.30*** | 1.42*** | 1.48*** | 1.30*** | 1.54*** | 1.33*** |
| 65+ |  | 1.48*** | 1.38*** | 1.31*** | 1.16 | 1.23** | 1.45*** | 1.16 | 1.39*** | 1.20 |
| Gender (male) |  |  |  |  |  |  |  |  |  |  |
| Female |  | 1.18*** | 1.18*** | 1.17*** | 1.12** | 1.10** | 1.10** | 1.06 | 1.12** | 1.05 |
| Other |  | 1.84*** | 1.79** | 1.81*** | 1.80** | 1.75*** | 1.78*** | 1.70** | 1.68** | 1.65** |
| Race (white) |  |  |  |  |  |  |  |  |  |  |
| Black |  |  | 0.89 | 0.88 | 0.87 | 0.80** | 0.90 | 0.85* | 0.86* | 0.85* |
| Hispanic |  |  | 0.85 | 0.86 | 0.87 | 0.80* | 0.86 | 0.85 | 0.82 | 0.84 |
| Asian |  |  | 0.75*** | 0.75*** | 0.78*** | 0.79*** | 0.74*** | 0.80*** | 0.80** | 0.81** |
| Other |  |  | 1.07 | 1.08 | 1.08 | 1.01 | 1.03 | 1.01 | 1.05 | 1.00 |
| Immigrant |  |  | 0.90 | 0.89 | 0.92 | 0.89 | 0.90 | 0.91 | 0.94 | 0.94 |
| Marital (married) |  |  |  |  |  |  |  |  |  |  |
| Previously married |  |  |  | 1.11 | 1.06 | 0.97 | 1.11 | 0.98 | 1.09 | 0.99 |
| Never married |  |  |  | 1.03 | 1.00 | 0.92 | 0.98 | 0.92 | 0.99 | 0.92 |
| Has children at home |  |  |  | 1.08 | 1.11* | 1.13** | 1.07 | 1.12** | 1.07 | 1.10* |
| Education (BA+) |  |  |  |  |  |  |  |  |  |  |
| HS or less |  |  |  |  |  |  |  |  | 1.39*** | 1.14* |
| Some postsecondary |  |  |  |  |  |  |  |  | 1.42*** | 1.20*** |
| Associate or equivalent |  |  |  |  |  |  |  |  | 1.43*** | 1.27*** |
| Main activity (employed) |  |  |  |  |  |  |  |  |  |  |
| Retired |  |  |  |  | 1.22** |  |  | 1.25** |  | 1.25** |
| Unemployed |  |  |  |  | 1.23** |  |  | 0.97 |  | 0.95 |
| Disabled |  |  |  |  | 2.53*** |  |  | 2.11*** |  | 2.04*** |
| Other |  |  |  |  | 1.07 |  |  | 0.99 |  | 0.97 |
| Family income (\$0-29k) | | | | | | | | | | |
| \$30-59k | | | | | | 0.75*** | | 0.87** | | 0.89* |
| \$60-89k | | | | | | 0.64*** | | 0.78*** | | 0.82*** |
| \$90-149k | | | | | | 0.58*** | | 0.76*** | | 0.81*** |
| \$150k+ | | | | | | 0.51*** | | 0.68*** | | 0.74*** |
| Financial hardship (no) |  |  |  |  |  |  |  |  |  |  |
| Little/slight |  |  |  |  |  |  | 1.36*** | 1.34*** |  | 1.35*** |
| Some |  |  |  |  |  |  | 1.77*** | 1.62*** |  | 1.64*** |
| Serious |  |  |  |  |  |  | 2.43*** | 2.17*** |  | 2.17*** |

\*p<.05, \*\*p<.01, \*\*\*p<.001
Regression models weighted using sampling weights; IRR shown (exponentiated coefficients). Pain is a continuous variable with 0-55 range. N=4,162.

**Table S3.** Tests of joint significance for all states/provinces/subnational units

|  |  |  |  |
| --- | --- | --- | --- |
|  | U.S. states | Canadian provinces | All subnational units |
| Null model | 44.7 (49) | 5.4 (10) | 82.8 (60)* |
| Model 1 | 44.8 (49) | 5.9 (20) | 77.1 (60)† |
| Model 2 | 49.0 (49) | 6.4 (10) | 68.4 (60) |
| Model 3 | 49.7 (49) | 6.8 (10) | 62.9 (60) |

\* $p < .05$ † $p < .1$
The table shows the value of the likelihood ratio test statistic and associated degrees of freedom from a chi-squared test of joint significant of the subnational units in four different negative binomial regression models of pain score.
The null model includes no covariates. Model 1 includes age and gender only. Model 2 includes age, gender, marital status, and children. Model 3 includes all covariates, that is, age, gender, marital status, children, education, main activity, family income, and financial hardship.

**Supplemental Table S4.**
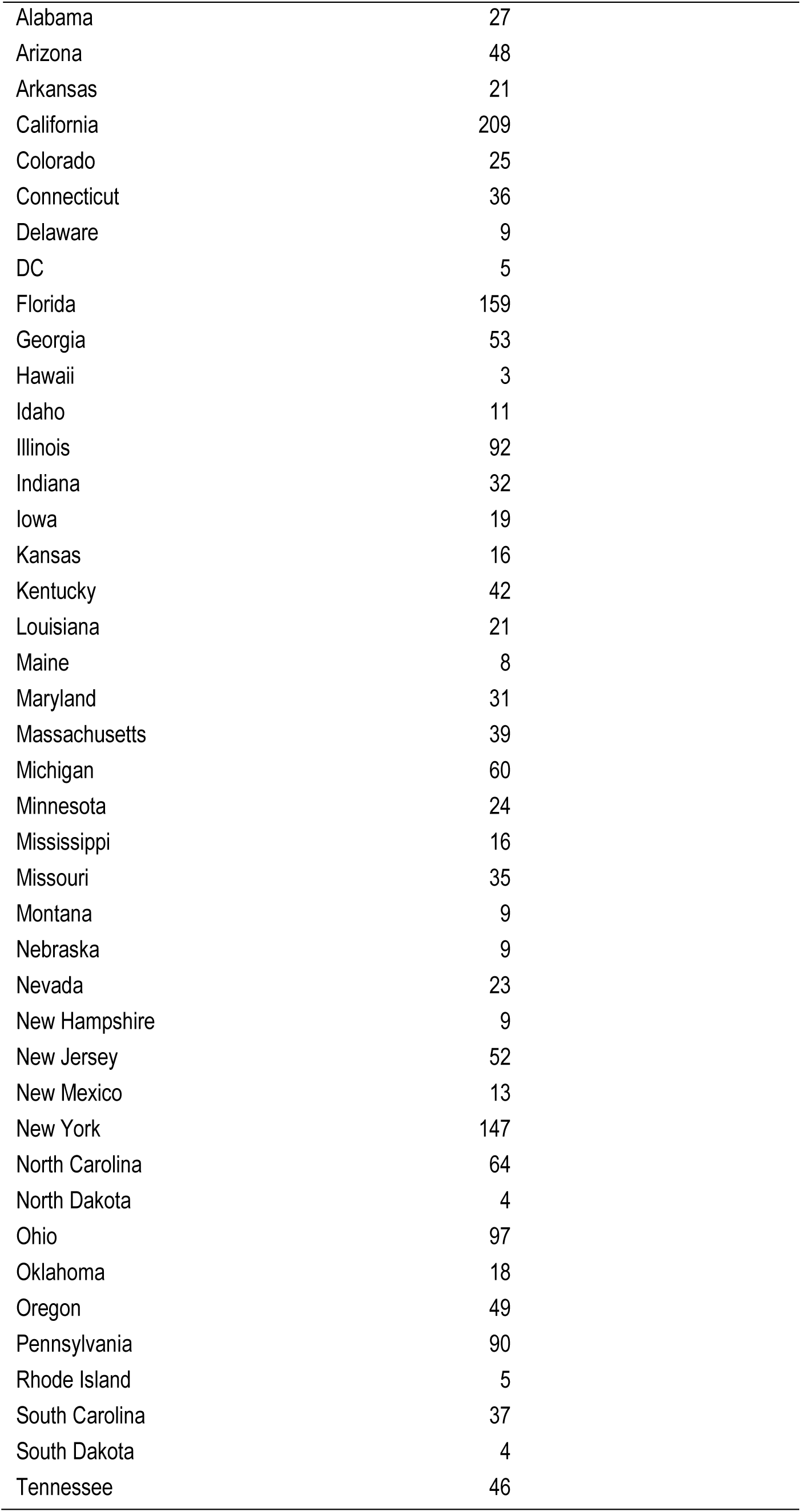
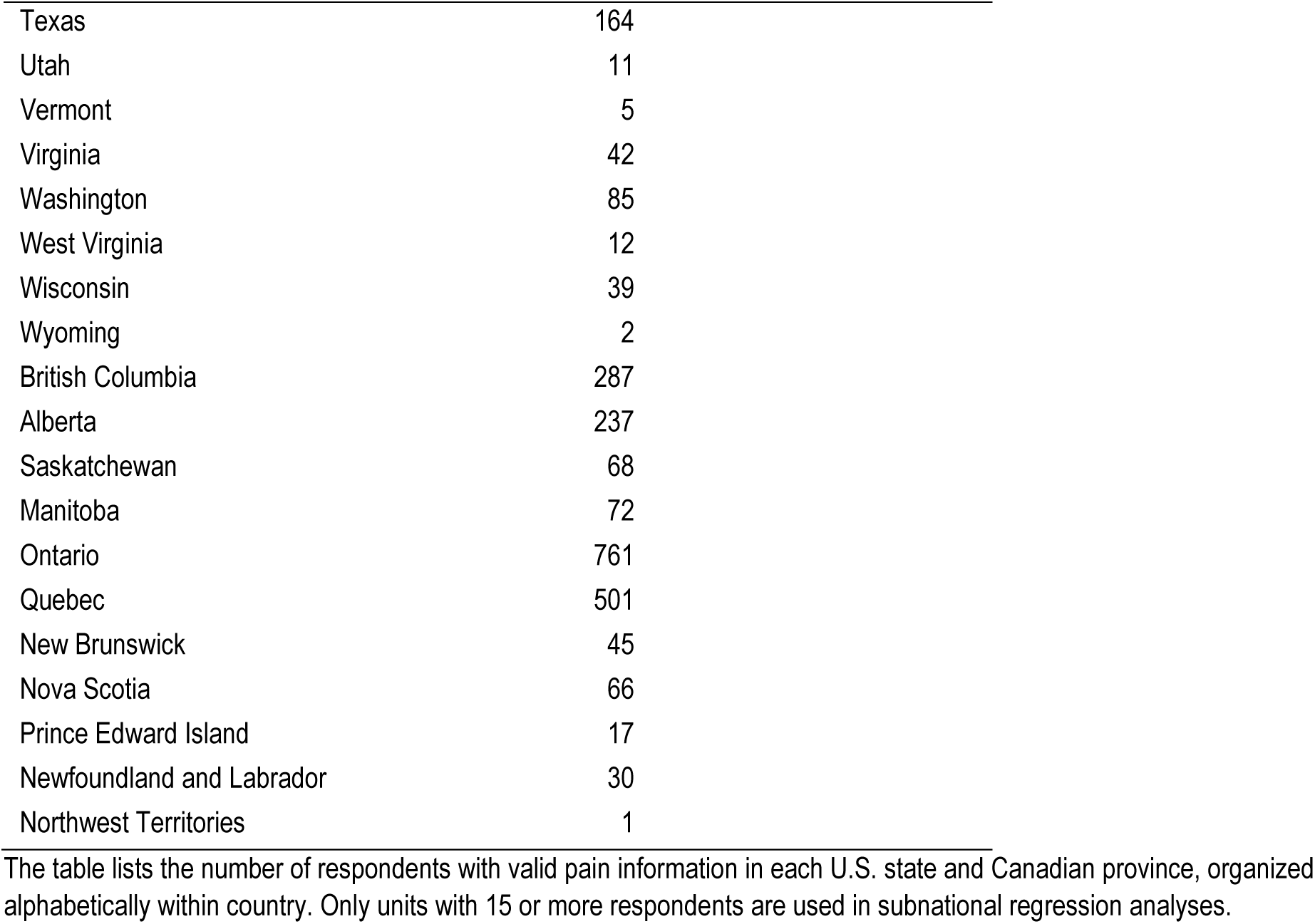
Number of respondents in each state and province (N=4,162)

|  |  |
| --- | --- |
| Alabama | 27 |
| Arizona | 48 |
| Arkansas | 21 |
| California | 209 |
| Colorado | 25 |
| Connecticut | 36 |
| Delaware | 9 |
| DC | 5 |
| Florida | 159 |
| Georgia | 53 |
| Hawaii | 3 |
| Idaho | 11 |
| Illinois | 92 |
| Indiana | 32 |
| Iowa | 19 |
| Kansas | 16 |
| Kentucky | 42 |
| Louisiana | 21 |
| Maine | 8 |
| Maryland | 31 |
| Massachusetts | 39 |
| Michigan | 60 |
| Minnesota | 24 |
| Mississippi | 16 |
| Missouri | 35 |
| Montana | 9 |
| Nebraska | 9 |
| Nevada | 23 |
| New Hampshire | 9 |
| New Jersey | 52 |
| New Mexico | 13 |
| New York | 147 |
| North Carolina | 64 |
| North Dakota | 4 |
| Ohio | 97 |
| Oklahoma | 18 |
| Oregon | 49 |
| Pennsylvania | 90 |
| Rhode Island | 5 |
| South Carolina | 37 |
| South Dakota | 4 |
| Tennessee | 46 |
| Texas | 164 |
| Utah | 11 |
| Vermont | 5 |
| Virginia | 42 |
| Washington | 85 |
| West Virginia | 12 |
| Wisconsin | 39 |
| Wyoming | 2 |
| British Columbia | 287 |
| Alberta | 237 |
| Saskatchewan | 68 |
| Manitoba | 72 |
| Ontario | 761 |
| Quebec | 501 |
| New Brunswick | 45 |
| Nova Scotia | 66 |
| Prince Edward Island | 17 |
| Newfoundland and Labrador | 30 |
| Northwest Territories | 1 |

The table lists the number of respondents with valid pain information in each U.S. state and Canadian province, organized alphabetically within country. Only units with 15 or more respondents are used in subnational regression analyses.

**Supplemental Table S5.**
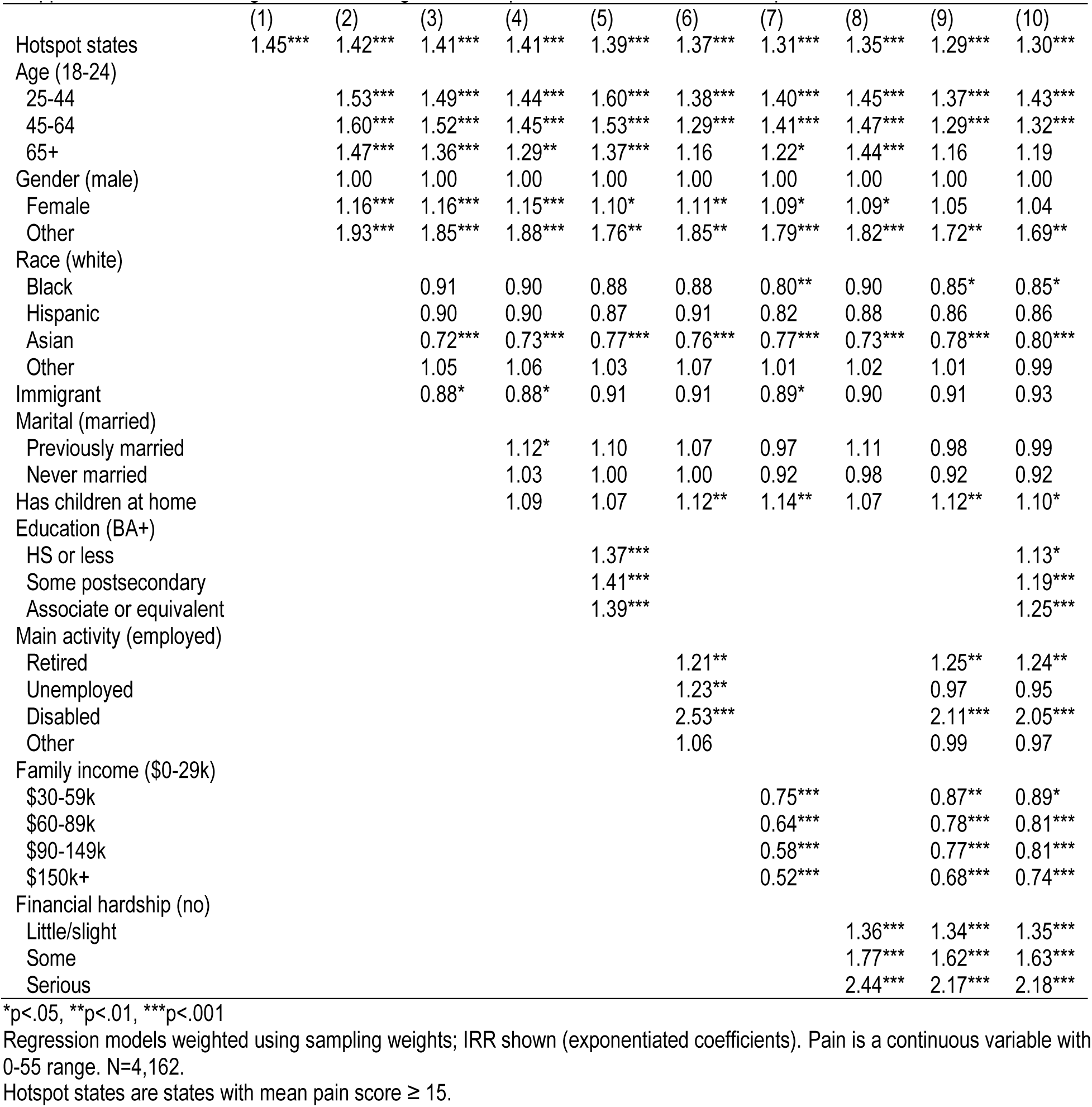
Negative binomial regressions of pain score as a function of hotspot areas, net of covariates.

